# The epidemiological characteristics of COVID-19 in Libya during the ongoing-armed conflict

**DOI:** 10.1101/2020.09.17.20196352

**Authors:** Mohamed Ali Daw, Abdallah Hussean El-Bouzedi, Mohamed Omar Ahmed, Ali Ali Alejenef

## Abstract

**Introductio*n*:** COVID-19 can have even more dire consequences in countries with ongoing armed conflict. Libya, the second largest African country, has been involved in a major conflict since 2011. This study analyzed the epidemiological situation of the COVID-19 pandemic in Libya, examined the impact of the armed conflict in Libya on the spread of the pandemic, and proposes strategies for dealing with the pandemic during this conflict.

**Methods:** We collected the available information on all COVID-19 cases in the different regions of Libya, covering the period from March 25 to May 25, 2020. The cumulative number of cases and the daily new cases are presented in a way to illustrate the patterns and trends of COVID-19 and the effect of the ongoing armed conflict was assessed regionally.

**Results:** A total of 698 cases of COVID-19 were reported in Libya during a period of three months. The number of cases varied from one region to another and was affected by the fighting. The largest number of cases was reported in the southern part of the country, which has been severely affected by the conflict in comparison to the eastern and western parts of the country.

**Conclusion:** This study describes the epidemiological pattern of COVID-19 in Libya and how it has been affected by the ongoing armed conflict. This conflict seems to have hindered access to populations and thereby masked the true dimensions of the pandemic. Hence, efforts should be combined to combat these consequences.

## Introduction

The COVID-19 pandemic has had major impacts on all aspects of life worldwide. No country can be considered safe, whether rich or poor. COVID-19 is a global concern not only as a huge health problem, but also socially, economically, politically and even ethically [1-3]. COVID-19 has caused huge numbers of deaths even in advanced, economically strong, politically stable countries with the best healthcare services [4,5].

COVID-19 has also affected most of the African countries, with over 103,875 cases and 3,184 deaths. The largest number of cases has been recorded in South Africa, and 64,388 cases have been reported in the WHO African region [6,7]. Many African countries suffer from internal conflicts complicated with the emergence of infectious diseases. One example is the Ebola epidemic in the Democratic Republic of Congo during armed conflict and geopolitical volatility, which led to the displacement of one million people [8,9]. Libya is the second largest African country, borders the Mediterranean Sea, and possesses huge natural resources. But the armed conflict that started in 2011 has continued and became more complicated by April 2019, resulting in extensive mortality, injury and population displacement [10,11]. Then, COVID-19 arrived

Armed conflicts cause deaths, injuries, destruction of infrastructures, and physical damage or destruction of healthcare delivery facilities. This facilitates the spread of newly emerging diseases. Hence, conflict zones are more susceptible to the spread of infectious diseases, including COVID-19. Concerns about the impact of the pandemic in countries experiencing armed conflict have been expressed. Such countries include Syria, Libya and Yemen, where the impact may go even beyond the borders of these countries [12,13].

Many studies have been published on all aspects of COVID-19, but not enough attention has been paid to its epidemiology in conflict zones [14,15], such as Libya, Sudan and Somalia. This study aimed to examine the status and patterns of COVID-19 in Libya during war time and the effect of the fighting on its emergence and spread. The study also seeks to highlight potential strategies to minimize the impact of this pandemic on Libyans during the conflict.

## Methods

The daily number of new COVID-19 cases was obtained from records made publicly available by national, provincial and municipal authorities throughout Libya and was recorded nationally and regionally. Information on the armed conflict was obtained from the army press offices in Tripoli and Benghazi and from the Ministry of State for Families of Martyrs, Injuries and Missing Persons, as described previously [10,11]. These data include the number of deaths and injuries. The epidemiological data of all confirmed COVID-19 patients from March 25, 2020 to June 25, 2020 in the centers for COVID-19 diagnosis were collected from all the Libyan regions. Data such as the geographic locations and population densities in the counties and regions are shown in supplement-1 [16].

Laboratory confirmation of COVID-19 in Libya is being done by the National Center for Disease Control. Nasopharyngeal and oropharyngeal swab samples are collected following standard safety procedures. RNA is extracted using QIAamp™ viral RNA mini kit from Qiagen™ according to the manufacturer’s instructions, as previously published [17]. Analysis is done by the real-time reverse transcriptase-polymerase chain reaction (RT-PCR) for all suspected cases following the protocol established by the WHO [18]. Biosafety cabinets are used and the work is done according to laboratory biosafety guidelines.

The geographic distribution of COVID-19 and the mapping of the armed conflict during the pandemic were determined as described [10,19,20]. As the armed combat can affect the prevalence of COVID-19 in the surrounding communities, the prevalence of COVID-19 in each city was recorded along with the distance to the fighting on the front lines. Distant cities were defined as those over 100 km away from the fighting area and adjacent cities as those within 100 km. A map of the regional distribution of COVID-19 from March 25 to June 25 in Libya was made, and the number of confirmed COVID-19 cases was color coded on the map.

Microsoft Excel and SPSS version 12.0 were used for data entry and analysis. The continuous variables included the daily cumulative number of cases, the daily number of newly confirmed cases, the number of deaths, and the numbers of severe and recovered cases [21]. The data were summarized as means ± standard deviation or medians, and the percentage of patients in each group was calculated. The available data were plotted to show the daily number of cumulative cases for each region and province. The geographical distribution of COVID-19 and the status of the war activities were drawn on a map of Libya.

## Results

The population in this study consisted of 698 confirmed COVID-19 cases reported from all over the Libyan regions (Table 1). Patient age ranged from 12 to 74 years, with an average of 43 years. Most of the patients (581, 83.2%) were males. Mortality was low (18, 2.6%). There were 140 recoveries (20.1%), leaving 540 cases still active (77.4%). Of the reported cases, 209 (29.9%) cases were initially imported and the rest were acquired within the Libyan territories. Given that the southern region has the smallest population, COVID-19 prevalence was highest in the south (292, 41.8 % of all cases). There were 235 cases (33.7%) in the western region, which is the most populous, and 171 cases (24.5%) in the eastern region.

**Table 1:** demographic features of the patients infected with COVID-19 in Libya

|  |  |  |  |  |
| --- | --- | --- | --- | --- |
| **Variables** | **Total no (%)** | **Death** | **Recovered** | **Active** |
|  | 698 | 18 (2.8) | 140(20.1) | 450(77.4) |
| **Age (years)** |  |  |  |  |
| ≤ 18 | 31 (4.4) | 2 (6.5) | 19 (61.3) | 10 (32.3) |
| 19-40 | 268 (38.4) |  | 38 (14.2) | 226 (84.3) |
| 41-65 | 327 (46.9) | 5 (1.3) | 65 (17.5) | 257 (69.1) |
| ≥ 66 | 72 (14.3) | 7 (9.7) | 18 (25) | 47 (65.3) |
| **Gender** |  |  |  |  |
| Male | 581 (83.2) | 13 (3.1) | 53 (9.1) | 515 (88.4) |
| Female | 117 (16.8) | 5 (4.3) | 87 (74.4) | 25 (21.4) |
| **Origin of cases** |  |  |  |  |
| Imported cases | 209 (29.9) | 7 (3.3) | 97 (46.4) | 105(50.2) |
| Local cases | 489 (70.1) | 11 (2.2) | 43 (8.8) | 435 (89) |
| **Geographic Location** |  |  |  |  |
| Western Region | 235 (33.7) | 4 (1.7) | 67 (28.5) | 164 (89.8) |
| Eastern Region | 171 (24.5) | 3 (1.6) | 41 (24) | 127 (74.3) |
| Southern region | 292 (41.8) | 11 (3.8) | 32 (11) | 249 (85.3) |

Figure 1 shows the cumulative numbers of COVID-19 cases in the country from 25 March to 25 June, 2020. Figure 1A shows the total number of cases in the three regions of Libya. The total number of cases continued to increase with time in all parts of the country. Figure 2B shows the daily change in the number of new cases since March 25, 2020. The daily number of new cases in the three regions showed an increasing trend, particularly in the south. The number of daily cases peaked on May 27 and then declined slightly, particularly in the Western and Eastern regions.

**Figure 1:**
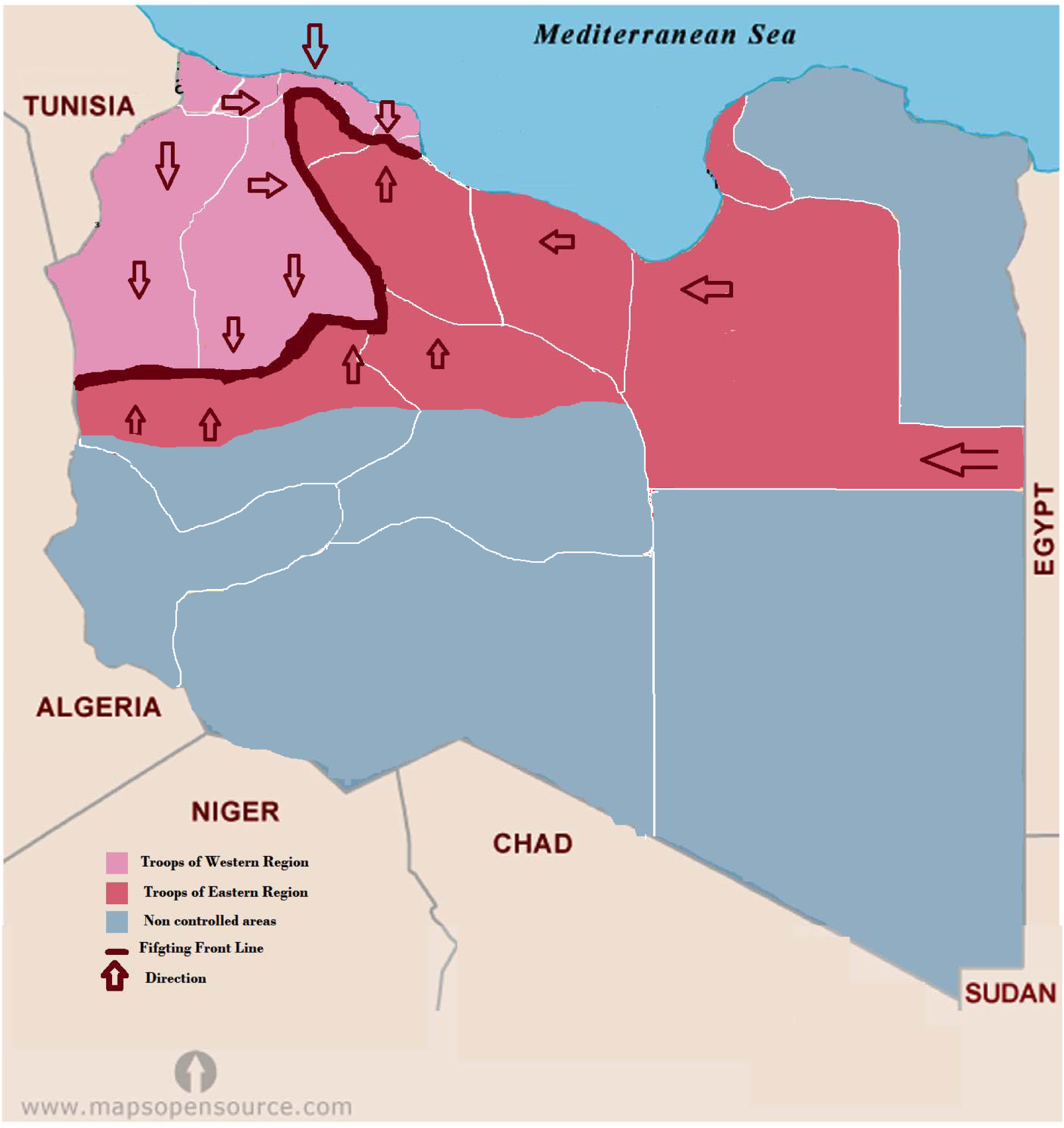
Map of Libya showing the geographic area controlled by each fighting group during the Emergence of COVID-19 in Libya.

**Figure 2:**
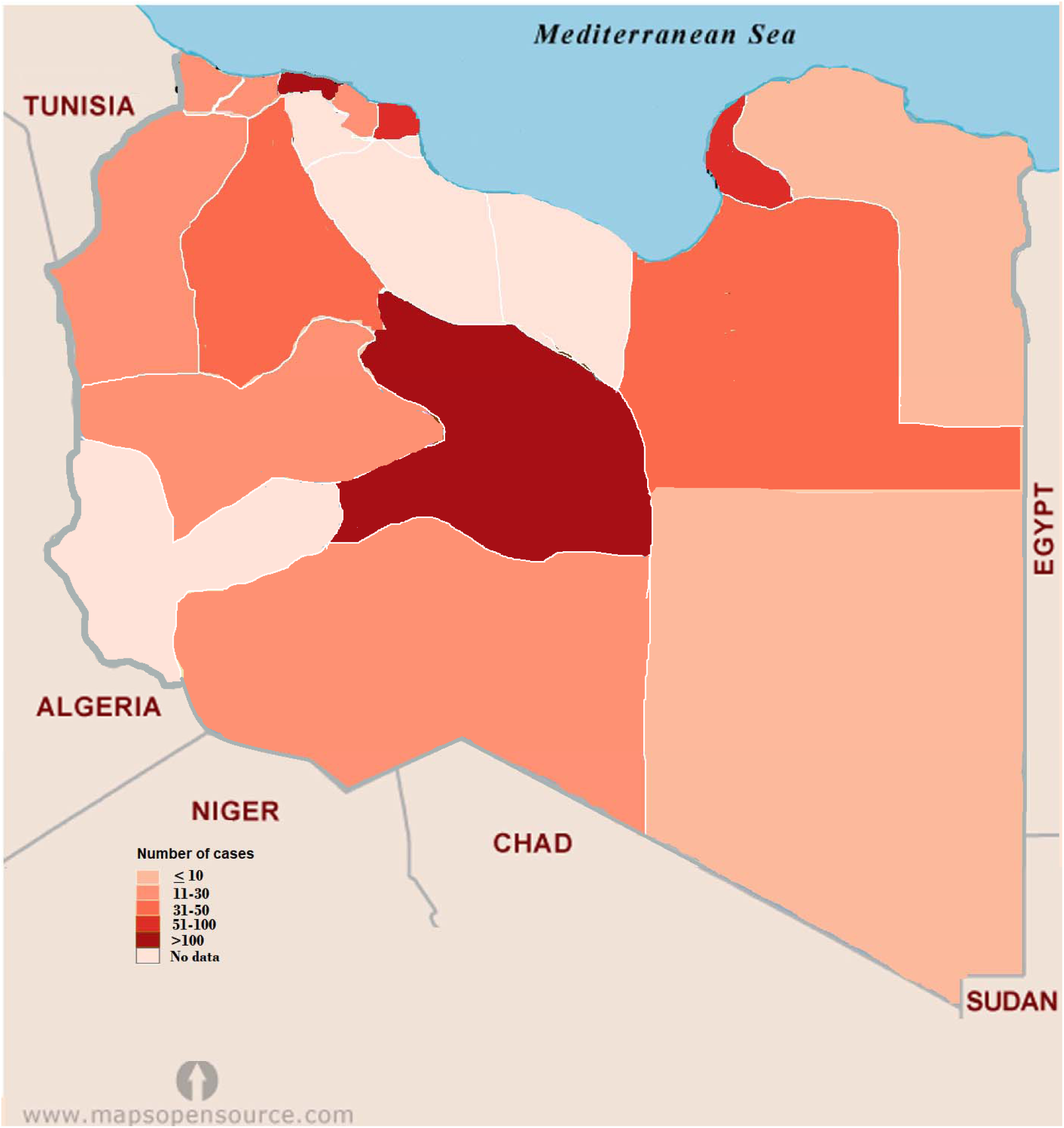
Geographic distribution of the reported cases of COVID-19 in Libya.

The geographic distribution of the cumulative cases of COVID-19 reported in Libya is shown in Figure 2. The number of cumulative cases is the daily sum of all confirmed cases from 25 March–25 June 2020. The regional distribution of COVID-19 is quite obvious, with significant spatial agglomeration. The southern region of Sebha had the largest number of cases (292, 41.8%), followed by Tripoli (110, 15.8%) and Musrata (87, 12.5%) in the west and Benghazi (99, 14.2%) in the east. In other Libyan counties such as Gharian, Ojula, Surman, Nalout, Ghat, and Jufra, the number of cases ranged from 10 (1.4%) to 79 (13.3%). No cases were reported in central counties such as Tarhona, Sert and Tawerga. This is evident that regional distribution of the disease varies significantly among the provinces and cities in Libya (*p* = 0.001).

Figure 3 shows the locations of the armed conflict during the emergence of COVID-19. Along the front lines, forces allied with the government located Tripoli (which controls mainly the western region) are pitted against forces allied with the government located in Benghazi (controlling the eastern region). The eastern forces tried to take control of the western region, which led to spread of the fighting from Jdabia till Tripoli. The cities which are within 100 km of the fighting are the most affected (*p* < 0.001). These include Sert, Tawerga and Tarhona, which have been devastated by the ongoing conflict, and in which no official health authority can work and no cases of COVID-19 have been reported. Distant counties located over 100 km away from the fighting, mainly the southern region, reported large numbers of infections.

**Figure 3:**
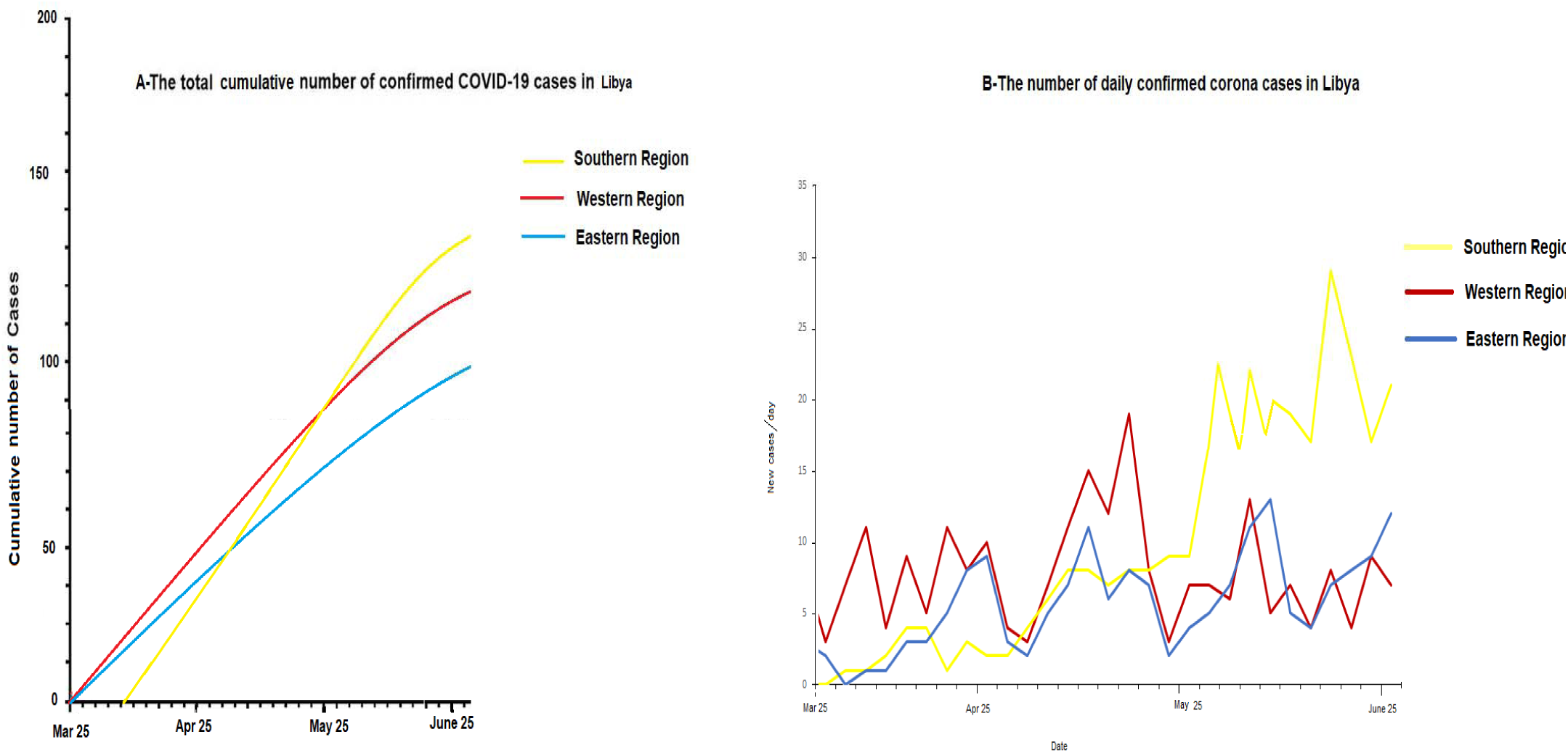
The graph shows the data of the number of confirmed cases in Libya between March 25, and June 25, 2020 (A). The cumulative number of COVID□19 cases in each Libyan region during the study period(B). The number of daily increased confirmed COVID-19 cases in each region.

## Discussion

Based on the total of 689 cases reported within a three-month period (March, 25 till June 25, 2020), we analyzed the epidemiological situation of COVID-19 all over Libya as well as the effect of the ongoing armed conflict on the pandemic patterns. The average age was 43 years and mainly males were affected, with a male-to-female ratio of 4.4:1.0. Of these patients, 540 (77.4%) are still hospitalized, 140 (20.1%) were discharged and 18 (2.6%) died. The number of daily new cases peaked between May 5 and May 7 in the western region, where the first cases were reported. Cases were reported in the eastern region on April 26, and later in the south on May 14. The total number of cases has started to increase substantially, particularly in the southern region and with no clear sign of declining. This suggests that the epidemic in Libya is not under control and that strict prevention and control measures have not been adopted. Nevertheless, despite the numerous challenges that the Libyan population has had to face since the armed conflict started in 2011, including deaths, injuries and internal displacement of populations, the response to the epidemic and the resilience of the healthcare system has been reasonable [22,23]. However, the situation remains precarious and a COVID-19 outbreak in this country would overload an already fragile healthcare system and poor baseline health status. Libya was the last country in the MENA region to report the first case of corona virus. However, preliminary epidemiological analysis carried out by Daw indicated COVID-19 might have arrived in Libya as early as January-February 2020, which has not been reported by the Libyan health authorities [24,25].

This study investigated the geographic distribution of COVID-19 in Libya and the effect of the ongoing armed conflict. The number of cases varied greatly from one region to another and the pattern was significantly influenced by the armed conflict. It is worth noting that Sebha in the southern region was the worst-hit city and had the highest number of infected cases. The increase in confirmed cases at any location will inevitably lead to increases in adjacent regions, a positive spillover effect. This was first seen after first cases were reported in Tripoli and Musrata in the western region, which were followed by spread to Zawia, Surman, Zletan and Alkomas. Likewise, in the eastern region the disease spread from Benghazi to Jalo, Ajdabia, Derna and Tubrak, and in the south from Sebha to Murzak, Obari, Wadishati and Ghat. This parallels, to a much smaller extent, the pandemic spread from Wuhan to the neighboring provinces and then all over China, and the Italian scenario, where the pandemic started in northern Italy, which at one time accounted for as much as 71.5% of the cases and 81.8% of the deaths, and then spread over the rest of Italy [26,27].

Comparison of the epidemiologic situations in different parts of Libya indicates that the ongoing armed conflict has affected the geographic spread of COVID-19 in two ways. On the one hand, it hindered access to populations and thus masked the actual status of the pandemic, particularly in cities such as Tarhona, Tawerga and Sert. On the other hand, it aggregated the spread of the pandemic to distant cities such as Sebha. Hence, the cross-national variation in the cumulative number of COVID-19 cases due the armed conflict is evident. Intervention strategies should be planned with that in mind [28, 29].

Controlling the emergence of infectious diseases in conflict situations is challenging because the fighting creates situations that facilitate the emergence of infectious diseases and enhance their transmission. These may include but are not limited to inadequate surveillance and response systems[30]. However, Libyan authorities have taken measures to cut off the source of infection, such as lockdown of cities and implementation of isolation procedures, but those were mainly restricted of Tripoli and Benghazi. Other regions were not comprehensively or effectively covered. Hence, mapping the disease enables the national authorities to ensure effective implementation of protective infectious disease interventions. This can be achieved by applying internationally accepted standards, guidelines and tools adapted to conflict situations, and this should be supported by specific training of health planners and health facility staff, and rapid mobilization of international experts to provide technical field support as required [30,31].

Despite the valuable epidemiological information that this study presents, it has several limitations and uncertainties, particularly as it was carried out in a conflict-ridden country where security is lacking and collecting accurate information is difficult. First, the number of reported cases is affected by uncertainties due to problems in accuracy in the daily reports of new notifications, particularly from the regions affected by ongoing armed conflict. Furthermore, it refers only to cases confirmed by molecular analysis, which is not feasible in all suspected situations. Daw[25] and Chen *et al* advocated that patients are the key cause of COVID-19 infection[32]. Accordingly, patients with mild or less severe symptoms should be taken into account as they may lead to an increase in infectivity and fatality. Second, the study did not analyze the medical care resources and healthcare capacities (number of hospital beds and physicians) used to combat the pandemic, especially that the patients are scattered over a very large area. The resources are most likely inadequate for a large influx of patients.

## Conclusion

This is one of the first studies to describe the epidemiological status of COVID-19 in an armed conflict area such as Libya and show the effect of the conflict on the epidemiologic pattern. As expected, COVID-19 spread from region to another, but this spread was influenced by the ongoing battles. By June 2020, the numbers of COVID□19 cases and deaths were still increasing in Libya, but the real situation might have been at least partially masked in communities in close proximity to battle zones and undetected in remote communities in the Sahara area. The ability to contain the spread of infections will depend on the development of enforceable national policies and success in effective education of the population in the reduction of infection risk according to international standards [33]. At the present time, achievement of such objectives is unpredictable if the ongoing armed conflict is not ended.

### What is already know on this topic

- This study is an epidemiological study carried on spread of COVID-19 in Libya, the second largest Country in Africa
- Armed conflict has an important impact of spread of COVID-19 in Libya
- National and regional cooperation should be combined to combat particularly within countries affected by conflicts

### What this study adds

- Highlights the epidemiological manifestations and the actual situation of COVID-19 in Libya; one of the largest countries in North Africa
- Illustrates the effect of the armed conflict on the spread of COVID-19 in Libya
- Contributes to discussion of ways to minimize the effects of armed conflicts on the spread of epidemics, particularly in Africa

## Data Availability

The data presented are solely available

## Authors’ contribution

M.D. conceived and designed the study. M.D., A.A. M.A & A.J. participated in the conduct of the study, and reviewed the manuscript. All authors approved the final manuscript.

## Acknowledgements

We are deeply grateful to the Department of Medical Microbiology& Immunology for their support and courage. Special thanks go to Mr. Ibrahem M Daw, Department of computer engineering, Faculty of Engineering, for his great efforts in mapping the results and Dr Amin Bredan (www.theeditor.be) for editing and correcting the manuscript.

